# Genome-wide association study of treatment resistant depression highlights shared biology with metabolic traits

**DOI:** 10.1101/2022.08.10.22278630

**Authors:** JooEun Kang, Victor M. Castro, Michael Ripperger, Sanan Venkatesh, David Burstein, Richard Karlsson Linnér, Daniel B. Rocha, Yirui Hu, Drew Wilimitis, Theodore Morley, Lide Han, Rachel Youngjung Kim, Yen-Chen Anne Feng, Tian Ge, Stephan Heckers, Georgios Voloudakis, Christopher Chabris, Panos Roussos, Thomas H McCoy, Colin G. Walsh, Roy H. Perlis, Douglas M. Ruderfer

**Author notes:** Corresponding authors: Clinical: Roy Perlis, Genomic: Douglas Ruderfer.

## Abstract

Treatment resistant depression (TRD), often defined by absence of symptomatic remission following at least two adequate treatment trials, occurs in roughly a third of all individuals with major depressive disorder (MDD). Prior work has suggested a significant common variant genetic component of liability to TRD, with heritability estimates of 8% when comparing to non-treatment resistant MDD. Despite this evidence of heritability, no replicated genetic loci have been identified and the genetic architecture of TRD remains unclear. A key barrier to this work has been the paucity of adequately powered cohorts for investigation, largely because of the challenge in prospectively investigating this phenotype. Using electroconvulsive therapy (ECT) as a surrogate for TRD, we applied standard machine learning methods to electronic health record (EHR) data to derive predicted probabilities of receiving ECT. We applied these probabilities as a quantitative trait in a genome-wide association study (GWAS) over 154,433 genotyped patients across four large biobanks. With this approach, we demonstrate heritability ranging from 2% to 4.2% and significant genetic overlap with cognition, attention deficit hyperactivity disorder, schizophrenia, alcohol and smoking traits and body mass index. We identify two genome-wide significant loci, both previously implicated in metabolic traits, suggesting shared biology and potential pharmacological implications. This work provides support for the utility of estimation of disease probability for genomic investigation and provides insights into the genetic architecture and biology of TRD.

## Introduction

Depression is a common, disabling mental illness, with lifetime prevalence estimates ranging from 6.6% to 21% globally^1^ and 16.9% in the United States^2^. Of the individuals with depression, more than 40% do not respond to 2 sequential antidepressant therapies and a third do not respond after 4 different treatments^3^. Treatment resistant depression (TRD) disproportionately accounts for the socioeconomic burden of depression, with over $25 billion spent annually in the United States^4^ and is associated with a significantly increased risk for suicide^5,6^. After decades of stasis, novel interventions for TRD have begun to emerge; however, such treatments remain costly and challenging to access, highlighting the need to better understand risk factors for this outcome^7,8^.

Despite decades of investigation^9^, the neurobiology of TRD is poorly understood. Prior studies have suggested a significant genetic component of TRD, with heritability estimates from common genetic variation ranging from 17% to 25% when compared to healthy controls^10,11^ and 8% when compared to non TRD MDD^11^. However, no identified genetic risk locus has been replicated in genome-wide association studies (GWAS). This likely reflects two key barriers to discovery: first, the challenges of attaining sufficient power for a heterogeneous phenotype and second, variable criteria for treatment responsiveness in defining TRD^12–15^. While the most common definition of TRD is a minimum of two prior treatment failures, there is limited consensus on the definition of treatment failure including exact measures of remission, length of adequate treatment trial duration, and adequate treatment dose^16^.

To address these challenges, we adopted two strategies. First, to increase power, we used large-scale clinical data to build risk prediction models where quantitative phenotypes can be generated for genetic samples in associated biobanks^17^. Secondly, to bypass the problems associated with categorizing treatment responsivity to specific medication classes or to specific number of antidepressant trials, we defined TRD on the basis of whether an individual with MDD had received a gold standard treatment indicated for TRD, electroconvulsive therapy (ECT)^18^. Together, these approaches enable the large-scale genetic analyses needed to characterize the genetic architecture of TRD and improve our understanding of its biological etiology.

Specifically, using ECT as a surrogate for TRD, we applied prediction models to electronic health record (EHR) data to derive posterior probabilities of receiving ECT, as absolute numbers of ECT cases in individual health systems were modest. After internally and externally validating these models, we used the probabilities as quantitative phenotypes to perform GWAS on over 154,000 patients with MDD across four large biobanks. We quantified the genetic contribution to TRD and the genetic overlap with both psychiatric and non-psychiatric phenotypes and other definitions of TRD. Finally, we implicated specific loci associated to increased likelihood of having TRD and needing ECT.

## Methods

### Study Settings

Clinical and genetic data were used from the EHRs and biobanks of Mass General Brigham (MGB), Vanderbilt University Medical Center (VUMC), Geisinger Health System (Geisinger; GHS), and the Million Veteran Program (MVP). MGB consists of 2 academic medical centers and 4 community and psychiatric hospitals in Eastern Massachusetts that serve over 6.5 million patients, and electronic health data were extracted from the Mass General Brigham Research Patient Data Registry (RPDR)^19^ and the Enterprise Data Warehouse. VUMC is an academic medical center in Nashville, Tennessee that manages over 2 million patient visits across Tennessee and its neighboring states each year. Its deidentified clinical EHR data is stored in the Synthetic Derivative (SD)^20^. Geisinger Health System is an academic medical center in Danville Pennsylvania and serve over 3 million patients in Pennsylvania. Deidentified electronic health data for consenting patients is extracted and stored by the Geisinger MyCode Community Health Initiative^21^. The Million Veteran Program^22^ study is based on the largest integrated health care system in the United States (Veterans Health Administration; 9 million people) and has over 825,000 US veteran participants with genetic information available for over 650,000 individuals.

### Clinical prediction model of TRD

We extracted de-identified clinical data of individuals with current ages of 18-90 years from the VUMC SD and MGB RPDR (**Figure 1A**). Individuals with MDD were identified using International Classification of Diseases, version 9 (ICD-9) codes 311.* (depression not otherwise specified), 296.2* (depressive episode), 296.3* (recurrent depression), and 300.4 (dysthymic disorder) and ICD-10 codes F32.** (depressive episode), F33.** (recurrent depressive disorder), and F34.1 (dysthymic disorder) with * as wildcard digits 0-9. Individuals with one or more ICD-9 or ICD-10 codes for bipolar disorders, schizophrenia, and psychotic disorders were excluded from analyses. Of the remaining individuals, TRD cases were defined using the Current Procedural Terminology (CPT) code for ECT (90870).

**Figure 1:**
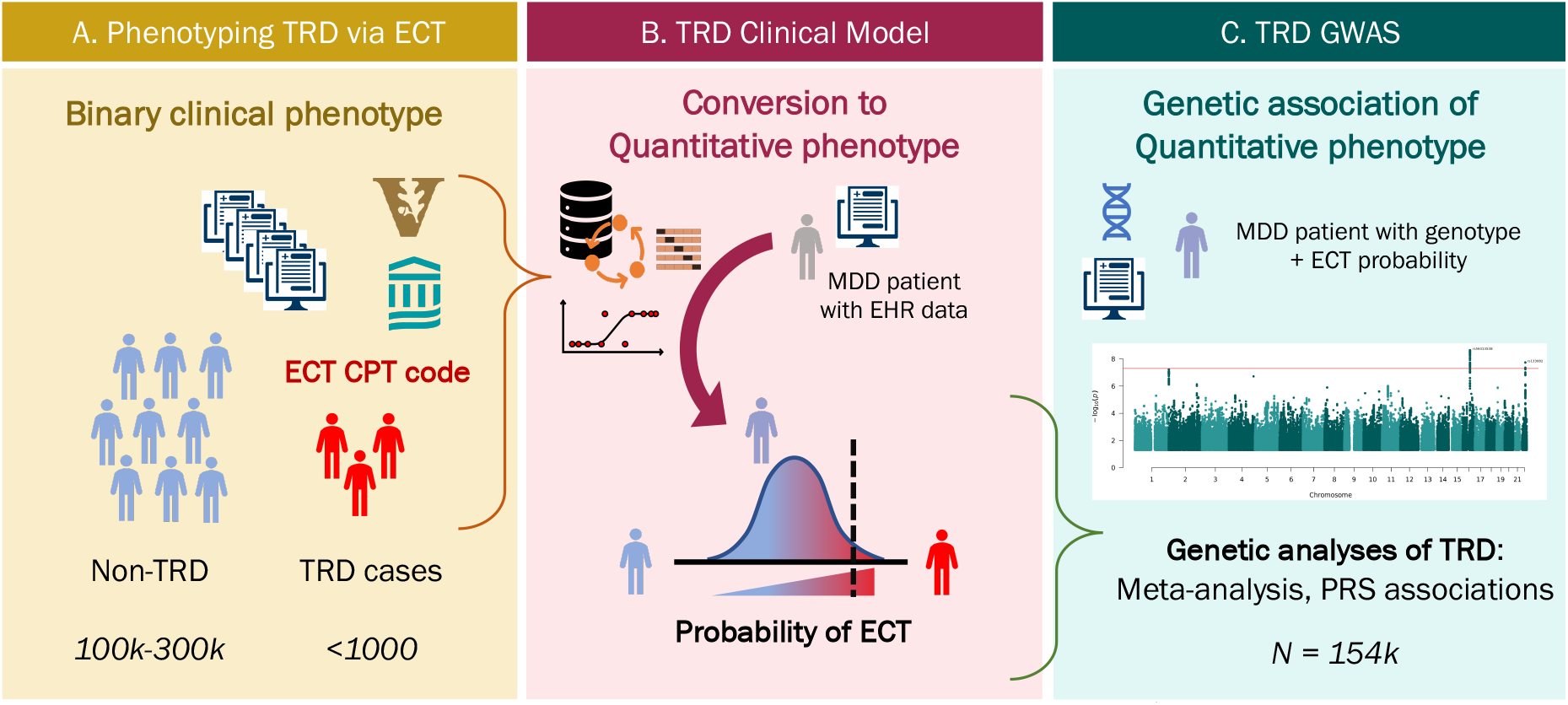
Schematic of the TRD clinical model generation and the genome-wide association study of the quantitative ECT prediction scores. A. TRD cases and non-TRD MDD controls were extracted from EHRs at VUMC and MGB B. Prediction models leveraging clinical data from the EHR were trained separately at VUMC and MGB and applied to 4 independent data sets to generate probabilities C. Probabilities were used as quantitative TRD phenotypes to perform two GWAS of TRD in 154,443 individuals.

Structured clinical data were included as predictors for the clinical model, including: demographics (age in years, categorical sex [Male, Female, Unknown], categorical race [White, Black, Asian, Hispanic, Other]), area deprivation index (ADI), diagnostic codes (log-transformed counts of historical Clinical Classification Software (CCS) counts)^23^, and medication (log-transformed counts of RxNorm-mapped ingredients). Of note, the VUMC ADI uses six features from the American Community Survey on the census tract level^24^, while MGB ADI includes 21 socioeconomic factors from the census on the zip-code level^25^. All data starting 24 hours prior to ECT for cases or prior to the last depression code for controls were excluded (right-censored) to avoid including features that are surrogates for the outcome (for example, the possibility that particular laboratory studies or pre-anesthesia procedures would directly proxy ECT). A minimum of at least two unique visit dates over four weeks before censoring date was required for study inclusion. The VUMC dataset was split into training and test sets where the test sample was comprised of only patients in the biobank. The remaining sample was then randomly split into 80% for training and 20% for validation. In MGB, the dataset was randomly split into 80% for training and 20% for testing regardless of biobank status. A LASSO model^26^ was trained separately at each site using Glmnet^27^ and hyperparameters were trained via a 10-fold cross-validation on the training data set (**Figure 1B**).

Each prediction model was validated internally and then externally at the other partner site (**Figure 1B**). Both the MGB and VUMC TRD models were further validated at Geisinger and MVP. Model performance was evaluated with discrimination metrics: Area Under the Receiver Operating Characteristic (AUROC) and Area Under the Precision-Recall Curve (AUPRC). Each genotyped individual from VUMC, MGB, Geisinger, and MVP had two predicted probabilities of ECT from either the MGB or VUMC TRD model, and these probabilities were used as quantitative phenotypes for genetic association analyses (**Figure 1C**).

### Validating TRD status of ECT cases

We extracted evidence of TRD based on number of unique antidepressants and explicit provider-based documentation in clinical notes. Specifically, we extracted antidepressants defined by the Anatomical Therapeutic Chemical (ATC) Classification code N06A and counted total number of unique medications. The free text of clinical notes were searched for at least one exactly matching instance of the following 4 phrases “resistant depression,” “resistant MDD,” “refractory depression,” or “refractory MDD.” TRD cases.

### Phenome-wide association study (PheWAS)

ICD-9 and ICD-10 codes were mapped to phecodes using version 1.2^28^. Cases were defined as those having more than 2 instances of the same phecode. Controls were patients never having documentation of the phecode. Patients were excluded if they had 1 instance of the phecode or were a case from a predefined list of excluded phecodes. Logistic regression was performed with presence or absence of phecode as the outcome. All phecodes with more than 100 cases and at least 1 case having received ECT were analyzed. The PheWAS R package^29^ was used to visualize results.

### Genotyping quality control, imputation and GWAS meta-analysis across four sites

Genotyping quality control (QC), imputation and GWAS were performed separately at each site. Standard QC protocols were applied removing variants based on high levels of genotype missingness or failing Hardy-Weinberg Equilibrium and removing individuals for excessive genotype missingness, high heterozygosity or sex discrepancies. Only individuals of European ancestries were retained as defined by the 1000 Genomes reference. Imputation and GWAS were performed using comparable reference panels and approaches. Site specific details are provided in the Supplementary Methods.

GWAS was conducted using linear regression of genotype on the VUMC and MGB TRD phenotypes in the individuals of European ancestries in 4 different clinical sites (VUMC, MGB, GHS, MVP) were meta-analyzed using inverse variance-weighted fixed effects model in METAL^30^. The weighted mean allele frequency was calculated weighted by the effective sample size per cohort. SNPs with a weighted minor allele frequency of < 1% or SNPs present in < 80% of total effective sample size were removed from the meta-analysis results.

### Heritability estimates and genetic correlation

LD score regression^31^ was used to estimate the phenotypic variance in TRD explained by common SNPs (SNP-heritability, 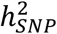) from GWAS summary statistics. 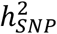 was calculated on the observed scale. LDSC bivariate genetic correlations attributable to genome-wide SNPs (rg) were estimated between GWAS of the TRD phenotypes and previously published GWAS of ECT^32^ or medication-defined TRD^11^, as well as other psychiatric and non-psychiatric risk factors. For previously noted epidemiological risk factors of TRD, the rg of TRD GWAS summary statistics with 29 other human diseases and traits was calculated using publicly available summary statistics (PMID listed in **Table S6**). The Bonferroni corrected significance threshold was P < 1.72×10^−3^, adjusting for 29 traits tested. Differences in rg between TRD_VUMC_ and TRD_MGB_ and differences in heritability between TRD meta-analyses before and after mtCOJO conditioning for BMI were tested for deviation from 0, using the block jackknife method, implemented in LDSC software^33^.

### Conditional GWAS using mtCOJO

The results of the GWAS of TRD were conditioned on the genetics of BMI using mtCOJO (multi-trait-based conditional & joint analysis using GWAS summary data)^34^, implemented in GCTA software^35^. mtCOJO estimates the effect size of a SNP on an outcome trait (eg. TRD) conditioned on exposure trait(s) (eg. BMI), using the genome-wide significant SNPs for the exposure trait as instruments to estimate the effect of the exposure on the outcome. It then performs a genome-wide conditioning of the estimated effect from the exposure, which provides conditioned effect sizes and P values for the outcome trait. We conditioned TRD on BMI, since higher BMI among TRD cases has been previously reported^11^. mtCOJO analysis was performed on the TRD meta-analyses as the outcome traits with the GIANT European ancestries GWAS summary statistics^36^ as the exposure trait since mtCOJO requires an ancestry-matched LD reference panel. In the selection of SNPs as instruments, independence was defined as SNPs more than 1 megabase (Mb) apart or with an LD r2 value < 0.05 based on the 1000 Genomes Project Phase 3 European reference panel^37^.

### Polygenic risk scoring (PRS)

PRS using the SNP weights from the quantitative TRD phenotype GWAS meta-analyses of the MGB or VUMC model were tested for association with ECT CPT code as well as the TRD prediction scores generated in independent target cohorts. The target cohorts were patients with MDD at VUMC and MGB. The meta-analysis of the TRD phenotypes was repeated excluding each cohort in turn to create independent discovery and target datasets. PRS analyses were performed using PRS-CS which places a continuous shrinkage prior on SNP effect sizes using a Bayesian regression framework^38^. The continuous shrinkage priors adapt the amount of shrinkage applied to each SNP to the strength of the associated GWAS signal based on the LD structure estimated from an external reference panel. Posterior SNP effects were generated in each cohort using PRS-CS and the 1000 Genomes European reference panel was used to estimate LD between SNPs. The PRS were calculated for each individual of the target cohort using Plink 1.9. PRS was tested for association with ECT cases vs control status in the target cohort using logistic regression model, covarying for PC1-PC10, sex, and age. PRS was also tested for association with MGB and VUMC TRD phenotypes using linear regression, covarying for PC1-PC10.

## Results

### Demographic and phenotypic characterization of patients with MDD receiving ECT across two healthcare systems

Leveraging longitudinal clinical data from EHRs at MGB and VUMC (see Methods), we identified 185,409 patients (MGB: 78,620, VUMC: 106,789) with a diagnostic code of MDD or depressive disorder. Depressive disorder was included as prior work in these health systems and others indicated that it is commonly applied by non-psychiatrists to capture MDD. We adopted this definition for consistency with numerous prior publications^8,39–41^ applying electronic health records to the study of major depressive episodes. Among those patients, 467 (MGB: 242, VUMC: 225) had at least one procedural code for ECT. The prevalence of ECT among individuals with MDD was 0.26% (MGB: 0.31%, VUMC: 0.21%) which is similar to the published prevalence of ECT of ∼0.25% among individuals with mood disorders^42^. The mean age at which VUMC cases received their first ECT CPT code was 53.8 ± 17.4 years, with a median ECT trial number of 15 (SD = 16); at MGB the mean age was 57 ± 17 years with a mean ECT trial number of 16 (SD = 19).

We identified multiple significant demographic differences between MDD patients receiving ECT and those who have not (**Table 1**). Consistent across the VUMC and MGB cohorts, ECT cases were 5 years older on average, 12% more likely to be male (although ECT was still more common in women), and 8.8% more likely to be white. However, while the VUMC ECT cases had lower mean body mass index (BMI) on average and at first ECT visit, BMI variables in the MGB cohort were comparable between cases and controls.

**Table 1:** Demographic characteristics of MGB and VUMC cohorts. In parentheses are percentages, standard deviations are reported after ±. Index time is 24 hours prior to first ECT for cases or last MDD code for controls. Age is defined as years between birth date and last EHR event. All non-listed races including unknown, and combinations are including in Other. BMI uses BMI values were cleaned for extreme outliers (> 80), unit mismatch and exclude individual measurements of age < 18. For ECT cases, the closest BMI measurement must have been within six months of the earliest ECT CPT code. Significance testing included t-test for quantitative values and two proportions Z-test for categorical variables.

| Demographic |  | VUMC patients |  |  | MGB patients |  |  |
| --- | --- | --- | --- | --- | --- | --- | --- |
|  |  | ECT case<br>(N=225) | MDD control<br>(N=106,564) | P-value | ECT case<br>(N=242) | MDD control<br>(N=78,378) | P-value |
| Age | Mean at earliest MDD | 51.2 ± 17.5 | 46.6 ± 19.3 | **1.23E-04** | 56.7 ± 16.3 | 51.2 ± 17.1 | **6.70E-07** |
|  | Mean at earliest ECT | 53.8 ± 17.4 |  |  | 57 ± 17 |  |  |
| Gender | Female | 121 (53.8%) | 70979 (66.6%) | **6.01E-05** | 145 (60%) | 55,334 (71%) | **0.002** |
|  | Male | 104 (46.2%) | 35548 (33.4%) | **6.00E-05** | 97 (40%) | 22,819 (29%) | **0.002** |
|  | Unknown | 0 (0%) | 1 (0%) | 1 | 0 (0%) | 3 (<0.1%) | 1 |
| Race | White | 200 (88.9%) | 88741 (83.3%) | **0.031** | 220 (91%) | 62,013 (79%) | **9.03E-06** |
|  | Black | 9 (4.0%) | 11552 (10.8%) | **1.41E-03** | 5 (2.1%) | 4,141 (5.3%) | **0.025** |
|  | Asian | 3 (1.3%) | 1064 (1.0%) | 0.87 | 5 (2.1%) | 1,664 (2.1%) | > 0.9 |
|  | Other | 13 (5.8%) | 5171 (4.8%) | 0.63 | 12 (5.0%) | 10,338 (13.2%) | **2.17E-04** |
| Ethnicity | Hispanic | 3 (1.3%) | 2189 (2.1%) | 0.60 | 4 (1.7%) | 6,883 (8.8%) | **1.38E-04** |
|  | Non-Hispanic | 213 (94.7%) | 99769 (93.7%) | 0.63 | 238 (98%) | 71,273 (91%) | **1.38E-04** |
|  | Unknown | 9 (4.0%) | 4570 (4.3%) | 0.96 | 0 (0%) | 0 (0%) | NA |
| BMI | Mean | 26.9 ± 6.81 | 28.2 ± 7.81 | **4.77E-04** | 28.1 ± 6.3 | 28.5 ± 6.6 | 0.4 |
|  | Closest to index date | 27.2 ± 6.81 | 29.8 ± 8.31 | **2.10E-07** | 28.0 ± 6.7 | 28.5 ± 6.8 | 0.3 |

### Validation of ECT cases as a TRD surrogate

Additional evidence of TRD status was extracted from the EHR using counts of unique antidepressants and explicit documentation by a provider of TRD in the patient’s clinical notes (see Methods). Among our ECT cases, 92% (VUMC) and 72% (MGB) had received at least two different antidepressants. On average, the ECT cases received twice as many antidepressants (VUMC: 4.9, MGB: 2.8 unique antidepressants) compared to the rest of the MDD cohort (VUMC: 2.2, MGB: 1.5 unique antidepressants). Of note, MGB prescriptions only included those written at MGB and did not include historical medications as included at VUMC. Filtering to written prescriptions at VUMC yielded 82% of ECT cases with at least two antidepressants suggesting that with historical prescriptions the proportion of MGB ECT cases with at least two antidepressants would be even higher.

Among the VUMC MDD cohort, 615 patients had direct documentation of TRD in their clinical record including 105 or 47% of the ECT cases representing a 182-fold increase of direct TRD documentation compared to the rest of the MDD cohort. The MGB dataset omits narrative clinical notes from McLean Hospital prior to 2017, which would tend to underestimate documentation of TRD given the greater proportion of inpatient care delivered there. Nevertheless, 266 MDD patients with documentation of TRD were identified including 41 or 15% of those with ECT which was a 58-fold increase of direct TRD documentation compared to the remaining MDD cohort.

In addition to gathering additional evidence of TRD among ECT cases, we also tested for phenotypes known to associate with TRD, such as increased suicidality and higher burden of comorbid psychiatric illnesses. We performed a phenome-wide association study (PheWAS) to test the relationship of ECT with other phenotypes (see Methods). PheWAS association results in the VUMC and MGB cohorts were highly correlated (r = 0.70. P = 4.19×10^−54^), with suicidality being the most significantly associated phenotype in both VUMC and MGB (VUMC: beta = 3.62, SE = 0.15, P = 2.67×10^−128^; MGB: beta = 2.57, SE = 0.18, P = 2.58×10^−46^). Other significantly associated phenotypes include psychiatric diseases like major depressive disorder, generalized anxiety disorder and other suicide-related traits (**Supplementary Figures 1-2**).

### Internal and external validation and generalizability of predicted TRD phenotypes

Models built to predict ECT separately at MGB and VUMC were tested both internally (within site) and externally (at different sites, **Figure 1**). Internal validation of the TRD models showed discrimination performance metrics on the held-out test sets at MGB and VUMC of AUROC 0.81 and 0.9, respectively, and AUPRC of 0.03 and 0.04, respectively (**Table 2**). External validation of the MGB TRD model on VUMC data showed AUROC of 0.83, AUPRC of 0.03 and applying the VUMC TRD model on MGB data showed AUROC of 0.83 and AUPRC of 0.03.

**Table 2:** Performance metrics of VUMC and MGB models across 4 sites. Test set is the independent sample both of the models were applied to. AUROC: area under the receiver operator curve; AUPRC: area under the precision recall curve. Bolded numbers are performance measures of internal validation.

| Site | ECT cases | Test Set |  | VUMC model |  | MGB model |  |
| --- | --- | --- | --- | --- | --- | --- | --- |
|  |  | Controls | Prevalence | AUROC | AUPRC | AUROC | AUPRC |
| VUMC | 59 | 33,248 | 0.18% | **0.90** | **0.036** | 0.83 | 0.030 |
| MGB | 207 | 43,520 | 0.47% | 0.83 | 0.030 | **0.91** | **0.080** |
| GHS | 353 | 190,841 | 0.18% | 0.84 | 0.021 | 0.78 | 0.023 |
| MVP | 600 | 259,925 | 0.23% | 0.81 | 0.024 | 0.81 | 0.040 |

To increase sample size and power for genetic analysis, features and weights of both models were applied to samples at two additional sites (**Table 2**), the Geisinger Health System (GHS: 353 cases, 190,841 controls) and the Million Veteran Program (MVP: 600 cases, 259,925 controls). Prediction performance remained consistently high for both models at GHS (VUMC model: AUROC: 0.84, AUPRC: 0.021; MGB model: AUROC: 0.78, AUPRC: 0.023) and MVP (VUMC model: AUROC: 0.81, AUPRC: 0.024; MGB model: AUROC: 0.81, AUPRC: 0.040). Features selected by LASSO with the highest weights included prescriptions of antipsychotics, diagnosis of mood disorders, and suicide in both models (**Table S1, Table S2**).

To assess the generalizability of our prediction models in capturing TRD in the absence of ECT, we tested the relationship between the prediction probabilities and the direct documentation of TRD in notes as defined earlier. Among the VUMC sample (where documentation was more comprehensive), patients with TRD mentions in their notes had 9-fold higher predictions probabilities with the model trained at VUMC (P = 1.14×10^−46^) and 7.2-fold with the model trained at MGB (P = 1.3×10^−50^). These increases in prediction probabilities remained high even after removing all ECT cases with 6.6-fold increase of prediction probabilities from the VUMC model (P = 3.3×10^−29^) and 6-fold increase from the MGB model (P = 2.9×10^−24^).

### TRD phenotypes show significant heritability and shared genetic architecture with each other

The posterior probabilities from the two TRD prediction models (VUMC and MGB) were rank normalized to generate two quantitative TRD phenotypes. Linear regressions of phenotype on genotype were performed separately limited to genotyped samples of European ancestries at VUMC (n = 15,305), MGB (n = 2,216), GHS (n = 39,353) and MVP (n = 97,649). Summary statistics from the four GWAS were then meta-analyzed across 154,433 samples using a variance-weighted fixed effect model. Significant heritability estimates of 0.042 (SE = 0.004, P = 4.90×10^−23^) for the MGB TRD meta-analysis (TRD_MGB_) and 0.020 (SE = 0.0036, P = 1.38×10^−8^) for the VUMC TRD meta-analysis (TRD_VUMC_) were estimated using LD-score regression^31^ (**Table 3**). Significant genetic correlation between TRD_VUMC_ and TRD_MGB_ was observed (rg = 0.66, SE 0.05, P = 1.3×10^−34^) (**Table S3**). The rg value reflects highly overlapping but non-identical phenotypes.

**Table 3:**
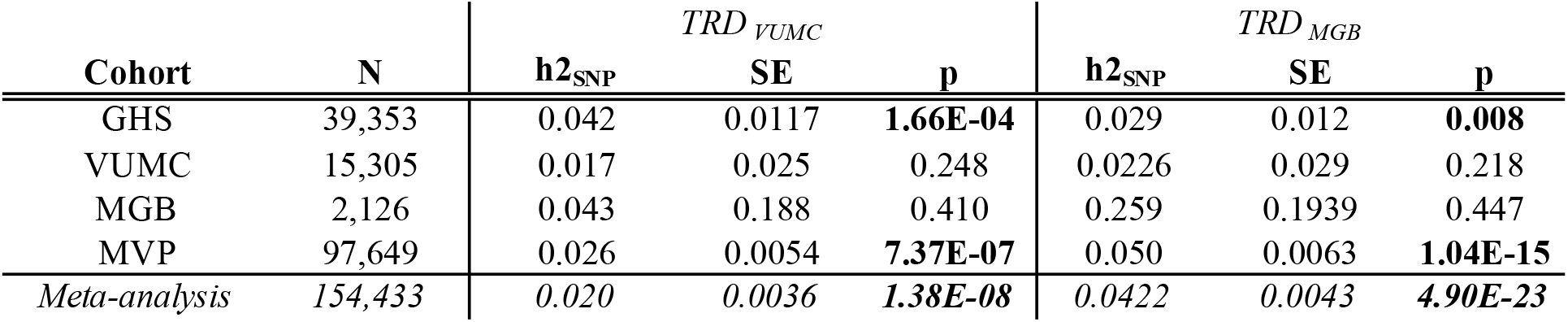
Heritability estimates of TRD GWAS within site and meta-analyses. Heritability was estimated with LD-score regression within each biobank site and the meta-analysis for both the VUMC and MGB TRD phenotypes.

| Cohort | N | *TRD<sub>VUMC</sub>* |  |  | *TRD<sub>MGB</sub>* |  |  |
| --- | --- | --- | --- | --- | --- | --- | --- |
|  |  | *h<sup>2</sup><sub>SNP</sub>* | SE | p | *h<sup>2</sup><sub>SNP</sub>* | SE | p |
| GHS | 39,353 | 0.042 | 0.0117 | **1.66E-04** | 0.029 | 0.012 | **0.008** |
| VUMC | 15,305 | 0.017 | 0.025 | 0.248 | 0.0226 | 0.029 | 0.218 |
| MGB | 2,126 | 0.043 | 0.188 | 0.410 | 0.259 | 0.1939 | 0.447 |
| MVP | 97,649 | 0.026 | 0.0054 | **7.37E-07** | 0.050 | 0.0063 | **1.04E-15** |
| *Meta-analysis* | *154,433* | *0.020* | *0.0036* | ***1.38E-08*** | *0.0422* | *0.0043* | ***4.90E-23*** |

We then examined the genetic correlation of our TRD meta-analyses with two prior GWAS of TRD (**Table S3**). The first defined TRD based on antidepressant prescriptions in the UK Biobank (TRD_UKB_)^11^ and the second used ECT but compared cases to healthy controls as opposed to those with non-TRD MDD (TRD_PREFECT_)^32^. No significant genetic correlation was observed between TRD_MGB_ or TRD_VUMC_ with either TRD_PREFECT_ (TRD_VUMC_: rg = 0.14, SE = 0.12, P = 0.23; TRD_MGB_: rg = 0.09, SE = 0.10, P = 0.31) or TRD_UKB_ (TRD_VUMC_: rg = 0.07, SE = 0.18, P = 0.65; TRD_MGB_: rg = -0.05, SE = 0.13, P = 0.69). TRD_UKB_ and TRD_PREFECT_ were significantly correlated with each other (rg = 0.75, SE = 0.24, P = 0.003). Further, genome-wide significant loci from the previously published TRD GWAS^32,43^ did not replicate at nominal significance in either of our TRD model meta-analyses (**Table S4)**.

### Two novel genome-wide significant loci associated to TRD

Two genome-wide significant loci were identified in TRD_MGB_ (**Figure 2**). The most significant locus was located on chromosome 16 in the intronic region of *FTO* (index SNP = rs56313538, beta for G allele = -0.0220, SE = 0.0037, MAF = 0.4, P = 4.3×10^−11^, Cochran’s Q: 0.35, I^2^ heterogeneity index = 9.53) (**Figure 3A, Table S5**). No significant association was observed in TRD_VUMC_ (beta = -0.003, SE = 0.0037, P = 0.37). The SNP is in high LD (R^2^ = 1.0) with SNP rs9939609 that is strongly associated with BMI^36^ (beta=0.075, SE=2.9×10^−3^, P = 1.95×10^−145^) and weight^44^ via its regulation of *IRX3* expression^45^. Therefore, this locus has an inverse effect on BMI and TRD.

**Figure 2:**
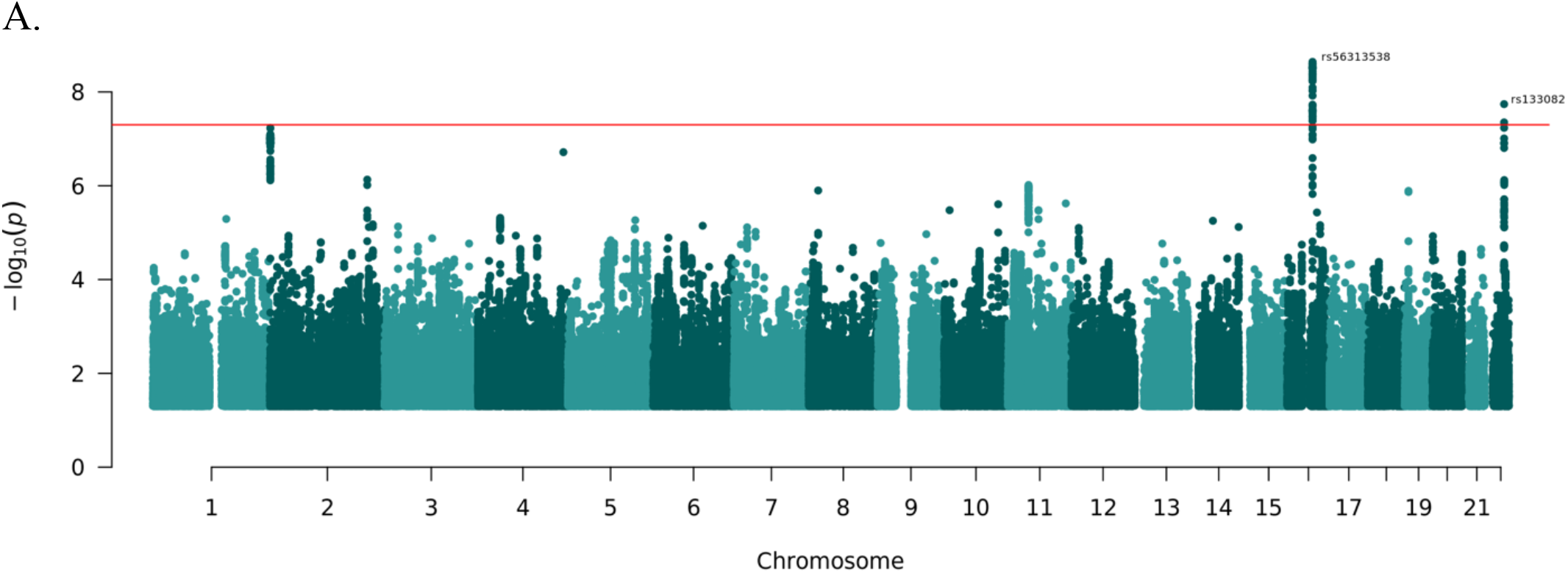
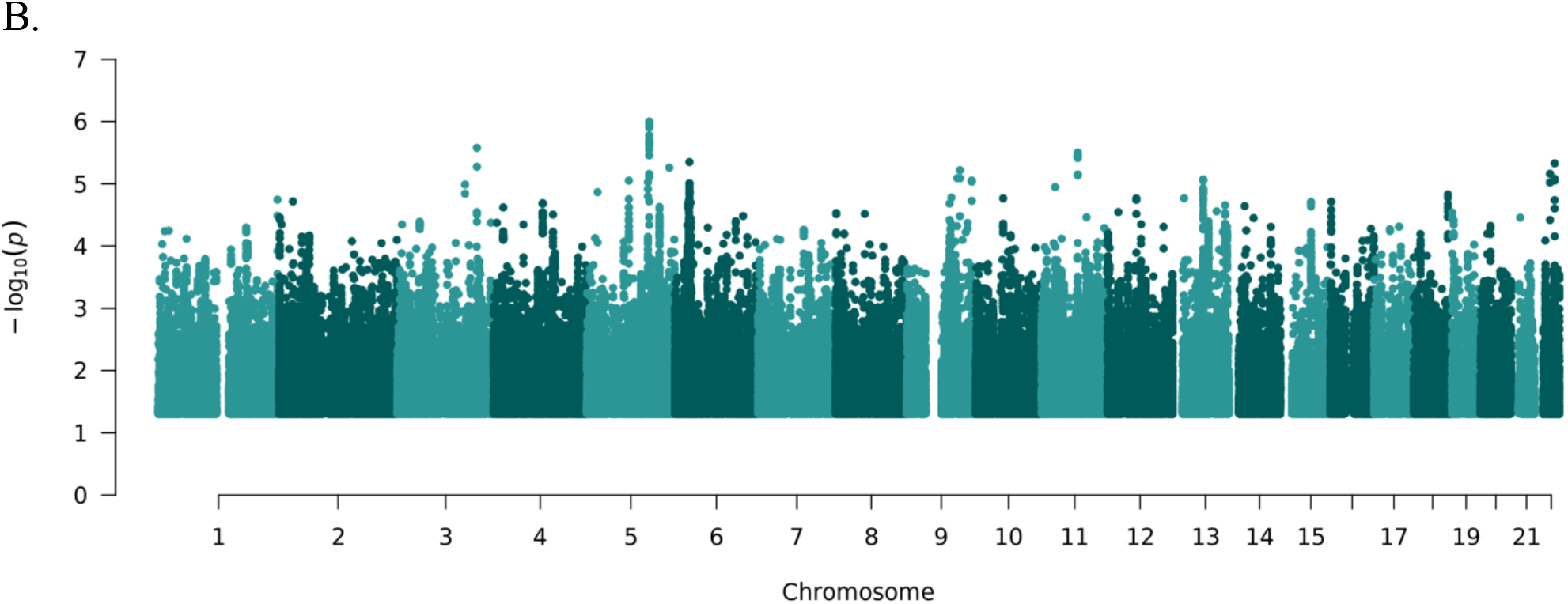
Manhattan plots of. A. MGB TRD meta-analysis (TRD_MGB_) and B. VUMC TRD meta-analysis (TRD_VUMC_)

**Figure 3:**
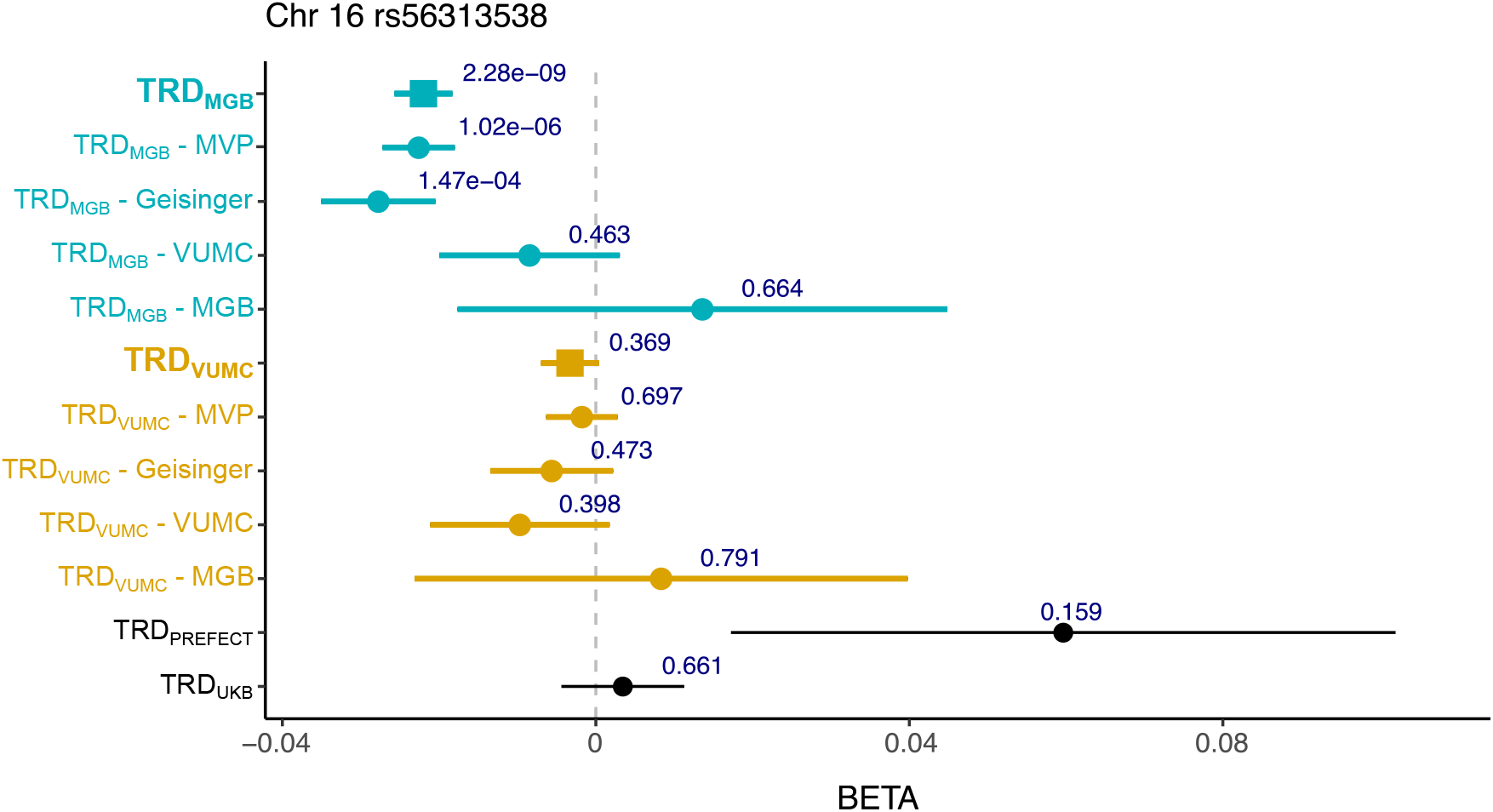
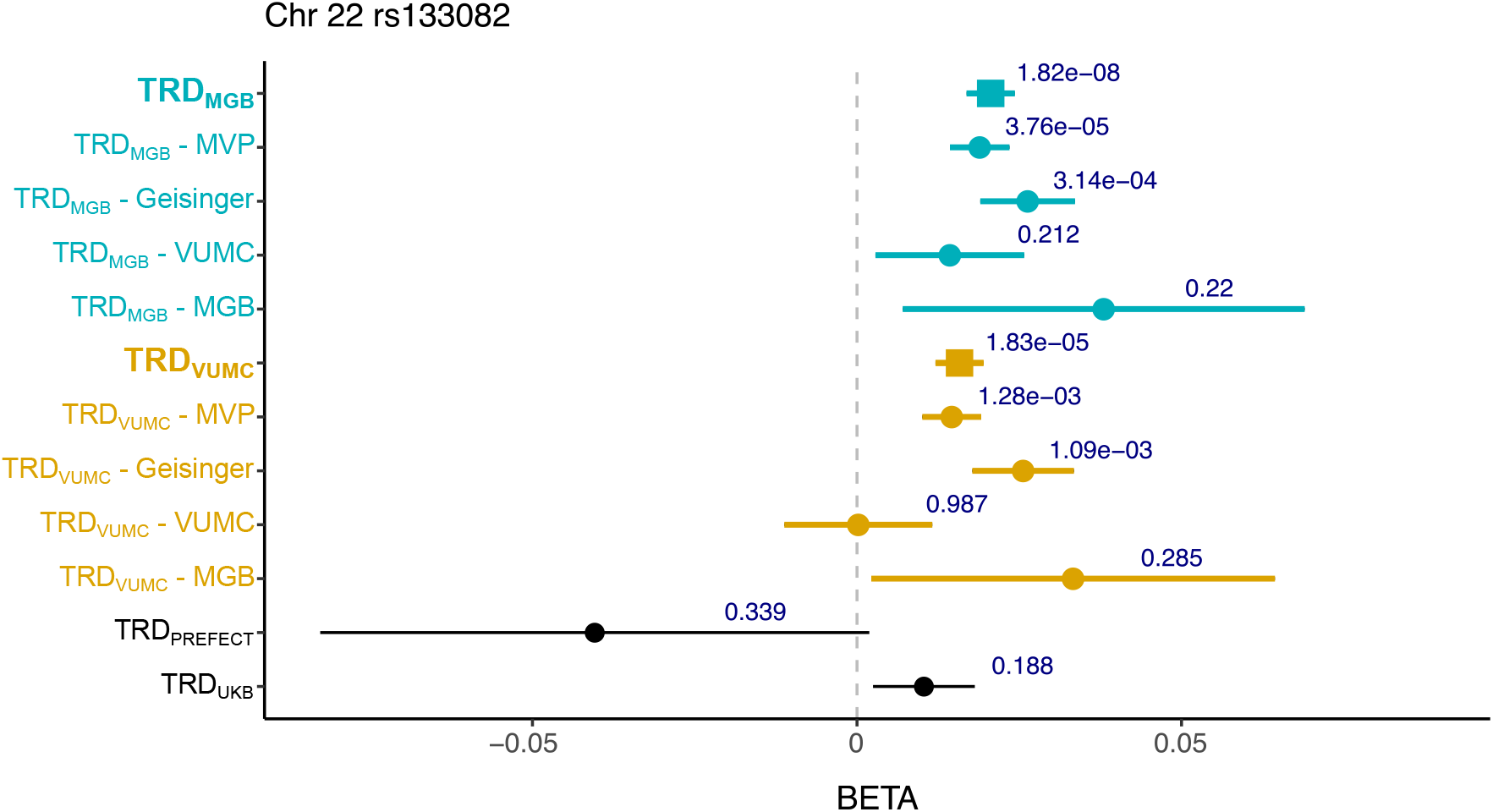
Forest plot of the genome-wide significant (GWS) locus rs8050136 on chromosome 16 and rs133082 on chromosome 22. The chromosome 16 loci is significantly associated with MGB TRD meta-analysis (TRD_MGB_) and the chromosome 22 loci is significantly associated with both MGB and VUMC TRD meta-analyses (TRD_VUMC_). Neither loci are significantly associated in two other TRD studies using different TRD definitions: GWAS of TRD based on antidepressant prescriptions in the UK Biobank^11^ (TRD_UKB_) and GWAS of TRD of ECT cases against healthy controls^48^ (TRD_PREFECT_). The points indicate the log odds ratio and the error bars show the standard error. The P value of association with each phenotype is shown above the error bars.

The second genome-wide significant locus was in an intergenic region on chromosome 22 (index SNP = rs133082, beta for C allele = 0.0206, SE = 0.0037, MAF = 0.44, P = 1.82×10^−8^, Cochran’s Q: 0.71, I^2^ heterogeneity index = 0) (**Figure 3B, Table S5**). Significant association was observed in TRD_VUMC_ although it did not surpass genome-wide correction (beta = 0.0158, SE = 0.0037, P = 1.83×10^−5^). This variant is significantly associated with decreased expression of the nearest gene, melanin concentrating hormone receptor 1 *(MCHR1*, 15kb away) in the dorsal lateral prefrontal cortex and increased expression in whole blood and T cells^46^. The variant is in high linkage disequilibrium (R^2^ = 0.76) with a genome-wide significant locus linked to increased risk of bipolar disorder in a recent large GWAS which implicated the same gene^47^. Neither locus is significantly associated in prior published TRD GWAS.

### TRD polygenic risk scores associate with TRD phenotypes

Polygenic risk scores are a standard approach to collapsing aggregated risk from genome-wide association studies^49^. We tested for association of polygenic risk scores generated using our TRD meta-analyses and our TRD phenotypes (prediction probabilities) in the VUMC or MGB samples after excluding them from the meta-analysis (i.e., always leaving out the target sample). Among VUMC patients, PRS generated from TRD_MGB_ was significantly associated with both VUMC and MGB TRD phenotypes (VUMC P = 2.15×10^−5^, MGB: P = 1.06×10^−11^) and TRD_VUMC_ PRS was significantly associated with the MGB TRD phenotype but not the VUMC TRD phenotype (MGB P = 6.01×10^−4^, VUMC P = 0.0619) (**Table 4**). Among the substantially smaller set of MGB patients, neither TRD PRS was significantly associated with either TRD phenotype.

**Table 4:** Polygenic risk score association results. PRS were generated using psychiatric traits and TRD meta-analyses as discovery GWAS and applied to TRD phenotypes based on models in the VUMC and MGB patient samples. P-values are bolded if surpassing the multiple-test correction threshold.

| Discovery GWAS | Target trait | VUMC patients ($n = 15,305$) | | | MGB patients ($n = 2,126$) | | |
| --- | --- | --- | --- | --- | --- | --- | --- |
|  |  | BETA | SE | P | BETA | SE | P |
| TRD<sub>VUMC</sub> | VUMC TRD phenotype | 0.0159 | 8.51E-03 | 0.0619 | -0.0026 | 2.36E-02 | 0.914 |
| TRD<sub>MGB</sub> |  | 0.0337 | 9.83E-03 | **6.01E-04** | -0.0182 | 2.40E-02 | 0.449 |
| Depression |  | 0.0330 | 8.05E-03 | **4.11E-05** | 2.54E-03 | 2.18E-02 | 0.907 |
| Bipolar Disorder |  | 0.0172 | 8.19E-03 | 0.036 | -0.0170 | 2.21E-02 | 0.443 |
| Schizophrenia |  | 0.0402 | 8.10E-03 | **7.24E-07** | -0.0051 | 2.21E-02 | 0.819 |
| TRD<sub>VUMC</sub> | MGB TRD phenotype | 0.0364 | 8.56E-03 | **2.19E-05** | 0.0138 | 2.38E-02 | 0.562 |
| TRD<sub>MGB</sub> |  | 0.0673 | 9.89E-03 | **1.06E-11** | 0.0100 | 2.42E-02 | 0.679 |
| Depression |  | 0.0212 | 8.11E-03 | 8.96E-03 | 0.0262 | 2.20E-02 | 0.233 |
| Bipolar Disorder |  | 0.0307 | 8.24E-03 | **1.99E-04** | 0.0282 | 2.23E-02 | 0.206 |
| Schizophrenia |  | 0.0473 | 8.16E-03 | **6.79E-09** | 1.56E-02 | 2.23E-02 | 0.484 |

We next looked at whether PRS derived from the most recent PGC summary statistics of relevant psychiatric traits including depression^39^, schizophrenia and bipolar disorder associated with our TRD phenotypes in VUMC or MGB patients. We identified that the depression PRS was significantly associated with the VUMC TRD phenotype (P = 4.11×10^−5^) and nominally associated with the MGB TRD phenotype (P = 8.96×10^−3^). Despite excluding patients with diagnoses of bipolar disorder or schizophrenia defined by at least one diagnostic code, we found that schizophrenia^50^ PRS was significantly associated with both the MGB and VUMC TRD phenotypes (**Table 4**, MGB: linear regression P = 6.79×10^−9^, VUMC: P = 7.24×10^−7^), and bipolar disorder^47^ PRS was significantly associated with MGB TRD phenotype (P = 1.99×10^−4^) and nominally associated with VUMC TRD phenotype (P=0.036). Among the smaller set of MDD patients in MGB, schizophrenia and bipolar disorder PRS were not significantly associated with either site’s TRD phenotype.

### Significant genetic overlap between TRD and psychiatric, cognitive, substance use and metabolic traits

To study the genetic overlap between TRD and psychiatric and non-psychiatric traits previously associated to TRD, genetic correlations were estimated. Both TRD meta-analyses showed significant positive genetic correlations, after multiple test correction, with cognitive traits including years of education (TRD_VUMC_: rg = 0.21; TRD_MGB_: rg = 0.46) and intelligence (TRD_VUMC_: rg = 0.20; TRD_MGB_: rg = 0.28), and significant negative genetic correlations with ADHD (TRD_VUMC_: rg = -0.27; TRD_MGB_: rg = -0.40), alcohol dependence (TRD_VUMC_: rg = -0.50; TRD_MGB_: rg = -0.38) and smoking traits (TRD_VUMC_: rg = -0.26; TRD_MGB_: rg = -0.42) (**Figure 4**). Both TRD meta-analyses also showed significant negative genetic correlations with BMI (TRD_VUMC_: rg = -0.33; TRD_MGB_: rg = -0.65). While the two TRD meta-analyses shared substantial genetic architecture, there were noticeable difference in genetic correlations across a subset of traits. Traits with significantly stronger genetic correlations in the MGB meta-analysis, based on a block jackknife approach in LD score regression^33^ included negative associations with BMI (P = 3.00×10^−11^) and type 2 diabetes (P = 4.67×10^−8^), and positive associations with educational attainment (P = 4.59×10^−9^) and marijuana use (P = 5.03×10^−6^). Traits that had a significantly stronger genetic correlation with the VUMC meta-analysis were neuroticism (P = 7.24×10^−5^), and multiple measures of alcohol use disorders, AUDIT-C (P = 3.54×10^−6^), and AUDIT-T (P = 3.67×10^−6^) (**Figure 4**).

**Figure 4:**
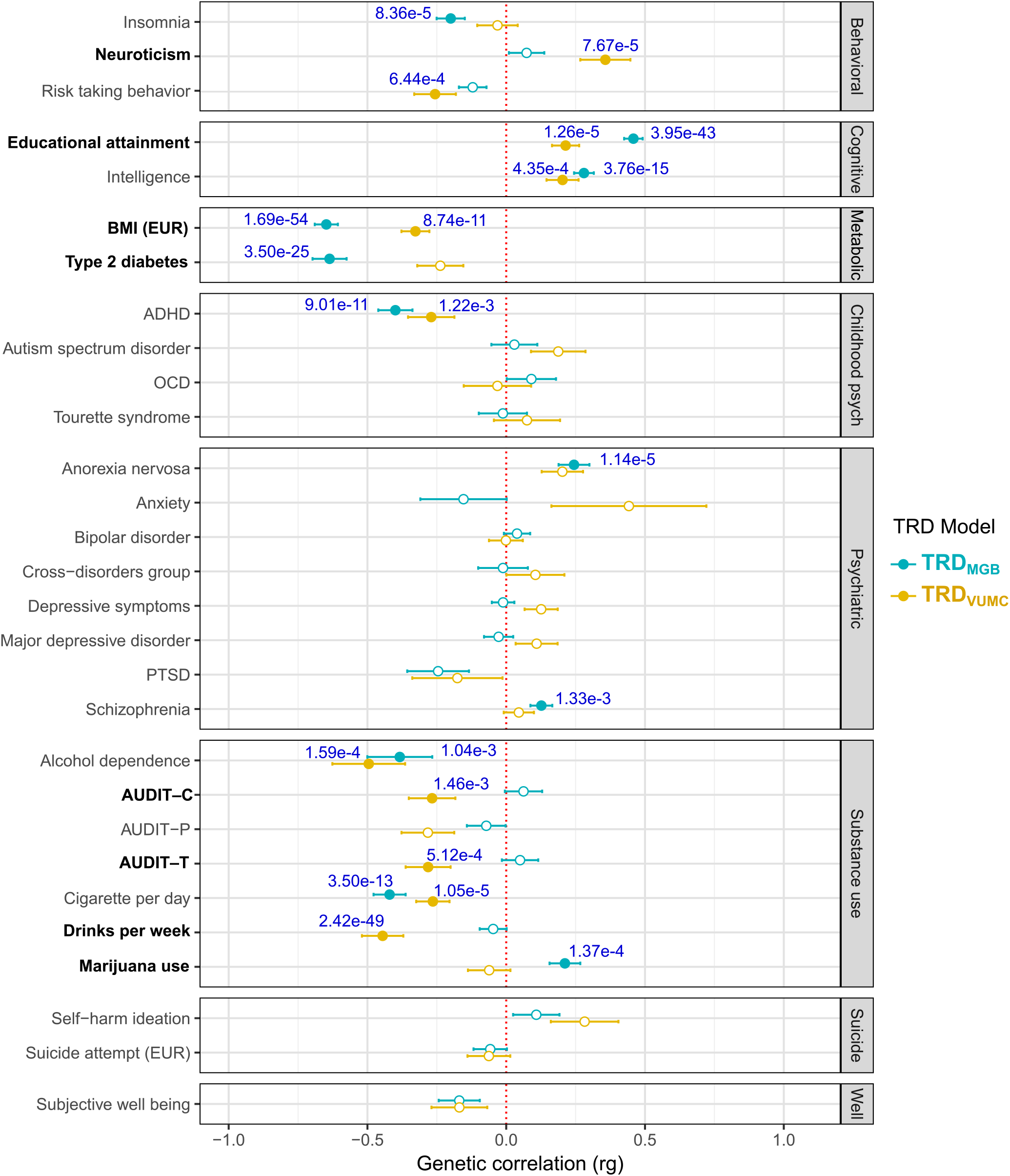
Genetic correlations of TRD_MGB_ and TRD_VUMC_ with psychiatric and non-psychiatric traits. Unfilled points indicate genetic correlations that did not pass the Bonferroni-corrected significance threshold P<1.72×10^−3^ (29 traits tested). Error bars represent the standard error. P values indicate significance after Bonferroni correction. Bolded traits show significant differences in genetic correlations between the two meta-analyses. Source GWAS of psychiatric and non-psychiatric traits are available in Table S6. BMI-body mass index, ADHD-attention-deficit/hyperactivity disorder, OCD-obsessive compulsive disorder, PTSD-post-traumatic stress disorder, AUDIT-C-Alcohol Use Disorders Identification Test-C (measure of quantity of alcohol consumption), AUDIT-P-measure of problematic consequences of drinking, AUDIT-T-total score of AUDIT.

Since we observed strong phenotypic and genetic association between TRD and BMI, we further examined the role that the genetic factors of BMI were having on our results by conditioning our TRD meta-analyses on the genetics architecture of BMI (see Methods). Significant differences in genetic correlations were confined to BMI for both meta-analyses and Type 2 diabetes for TRD_MGB_ only (**Supplementary Figures 3-4**). Among the identified loci, chromosome 16 is a known BMI locus and was used as an instrument SNP for the conditioning analysis so was removed. However, the conditioning on BMI had limited effect on the association statistics from the locus on chromosome 22 in MGB (beta = 0.0194, SE = 0.004, P = 2.02×10^−7^) and VUMC (beta = 0.0154, SE = 0.004, P = 3.27×10^−5^) (**Table S5**).

## Discussion

In this genetic study of TRD, we found low but significant heritability of TRD, with two novel genome-wide significant loci and significant genetic overlap with schizophrenia, cognitive and substance abuse traits, as well as BMI. Application of a computed phenotype from biobank-linked electronic health records allowed detection of these effects in a total of 154,433 individuals across 4 sites. Understanding the genetic architecture of TRD is important for quantifying the role of genetics in treatment response to move beyond decades-old pharmacogenomic studies. Further, identifying risk loci could facilitate efforts to identify novel treatments considering the modest response rates observed for interventions other than ECT.

Our data support that ECT use among MDD patients is overwhelmingly reserved for those with TRD. ECT remains a rare procedure with a prevalence among individuals with MDD of well below 1% due to stigma, side effects, access, socioeconomic and other factors. Even with 154,433 patients, a case-control approach comparing ECT cases to MDD controls would have been underpowered with ECT case numbers of only a couple thousand across all four clinical sites. Leveraging models that can predict ECT from large repositories of clinical data and assign probabilities as quantitative phenotypes allows for substantial increase in power in genetic studies. We showed that our ECT-based prediction models trained both at MGB and VUMC were robust to external validation across three independent healthcare systems. Importantly, our predicted phenotypes demonstrate generalizability in predicting TRD based on clinical documentation even in the absence of ECT.

With quantitative phenotypes, we increased power across the entire genotyped cohort of 154,433 patients which resulted in a significant SNP heritability of 2-4% and significant genetic correlation between the two TRD meta-analyses. Both TRD meta-analyses showed significant positive genetic overlap with cognitive traits, and significant negative genetic correlations with ADHD, alcohol and smoking traits, and BMI. We also saw modest evidence of genetic risk of schizophrenia and bipolar disorder even after removing patients with any diagnostic evidence of schizophrenia or bipolar disorder from our population, further implicating potential genetic risk of severe psychiatric disorders to likelihood of TRD. Despite the high genetic correlation across models, genetic overlap with other traits differed significantly representing potential differences in clinical population, general population and/or clinical decision making around ECT. We did not see any genetic correlation with other genetic studies of TRD. However, the comparable ECT studies used healthy controls and these two studies were highly genetically correlated with each other pointing to the potential that they are predominantly capturing depression genetic architecture as opposed to TRD genetic architecture. Our work shows there is a significant but small contribution of genetics to TRD as defined by ECT. Large studies are currently underway to collect tens of thousands of ECT cases for a case-control study^51^ and the comparison to this more timely and efficient approach will be important.

We discovered two genome-wide significant loci both with prior implications to metabolic traits. The most significant locus was in the intergenic region of the obesity and BMI-related *FTO* gene on chromosome 16. The index variant is in perfect linkage disequilibrium with the known BMI variant that regulates *IRX3* and *IRX5*. This locus was not supported by the TRD_VUMC_, consistent with MGB model having stronger genetic correlation with BMI. Of note, the *inverse* genetic association between TRD and BMI – that is, low BMI being associated with high risk of TRD – is supported by the positive correlation between TRD with anorexia nervosa, a metabopsychiatric illness characterized by extremely low body weights and a notorious lack of antidepressant responsivity in those with comorbid depression. Interestingly, BMI has previously been shown to moderate treatment response in patients with MDD receiving intravenous ketamine, where patients with higher BMI and obesity demonstrated a more robust acute antidepressant response to ketamine^52^. Our study is in line with such findings and extends this to incorporate the metabolic genetic vulnerabilities underlying differential antidepressant treatment responsivity.

The second risk gene *MCHR1*, encodes a G protein-coupled receptor protein linked to neuronal regulation of food intake as well as obesity and insulin resistance in mouse models^53^. This gene is highly expressed in the brain in regions that regulate body weight and appetite^54^ and has recently been implicated in bipolar disorder^47^, which is an illness associated with metabolic traits and severe depressive episodes. Moreover, inhibition of the *MCH1* signaling pathway results in anti-depressant and anorectic effects in murine models^55^, which is in line with our findings that implicate *MCHR1* in depression treatment responsivity phenotype along with association to lower BMI. This locus shares substantial support from the VUMC meta-analysis, and the effect size is largely constant even after conditioning on BMI. Together, our identification of *FTO* and *MCHR1* as genetic vulnerabilities for TRD supports a direct link between depression treatment responsivity and the complex metabolic regulatory pathways underlying energy balance including food intake and body weight homeostasis.

We note several limitations of our study, particularly the potential confounding of ECT population characteristics in our TRD clinical models. There were significant demographic differences between cases and controls where a typical ECT case tended to be an older, white, male with a lower mean BMI compared to MDD controls. These demographic differences could be driven by ascertainment in medical decisions leading to a patient receiving ECT or socioeconomic factors like access to a caregiver as patients need accompaniment after the inpatient ECT procedure. Demographic differences between the region around Nashville and eastern Massachusetts could also contribute to the differences observed in the VUMC and MGB TRD phenotypes, but the two meta-analyses showed significant genetic overlap (rg = 0.66), and both prediction models performed robustly in independent clinical sites with different demographics, especially in the Million Veteran Program cohort, which is significantly more male and older than the other cohorts. Phenotypes based on prediction models are not always representative of the original phenotype and could differ in important ways that modify genetic architecture and power. We were able to identify significant but low SNP-heritability (2-4%), meaning that even with our substantial improvements in power many more patients will be required to enable identification of additional genome-wide significant loci. We note that previous estimate of SNP-heritability of TRD within MDD patients using prescription data was only slightly higher at 8% but with a wider confidence interval (SE = 0.04)^11^.

Despite these limitations, this study supports the utility of investigating a validated proxy for TRD that can be readily extracted and predicted from electronic health records or administrative claims data. We confirm a significant but modest genetic contribution to TRD and provide insights into its overlap with other psychiatric and non-psychiatric phenotypes, in particular metabolic traits. This work lays the groundwork for future efforts to apply genomic data for biomarker and drug development in TRD.

## Data Availability

Summary statistics files and code used to generate data and figures are available in https://github.com/RuderferLab/TRD_GWAS.

## Acknowledgements

This work was funded by National Institutes of Mental Health grants R01MH116269 (DMR, CGW), R01MH121455 (DMR, CGW), R01MH116270 (DMR, CGW, RHP) and R01MH123804 (RHP), and National Institute of General Medical Sciences grant 5T32GM007347 (JK). The dataset(s) used for the analyses described were obtained from Vanderbilt University Medical Center’s BioVU, which is supported by institutional funding, private agencies and federal grants. These include the NIH-funded Shared Instrumentation Grant S10RR025141, and CTSA grants UL1TR002243, UL1TR000445 and UL1RR024975. Genomic data are also supported by investigator-led projects that include U01HG004798, R01NS032830, RC2GM092618, P50GM115305, U01HG006378, U19HL065962 and R01HD074711; and additional funding sources listed at https://victr.vanderbilt.edu/pub/biovu/. We thank Mass General Brigham Biobank for providing samples, genomic data, and health information data. This work was conducted in part using the resources of the Advanced Computing Center for Research and Education at Vanderbilt University, Nashville, TN. Genotyping data from the RGC-GHS DiscovEHR collaboration was generated by the Regeneron Genetics center.

This research is also based on data from the Million Veteran Program, Office of Research and Development, Veterans Health Administration, and was supported by award #MVP006. This publication does not represent the views of the Department of Veteran Affairs or the United States Government. This study was also supported by the National Institutes of Health (NIH), Bethesda, MD under award numbers K08MH122911 (GV), R01MH125246, R01AG067025, U01MH116442, R01MH109677 (PR), and by the Veterans Affairs Merit grants BX002395 and BX004189 (PR). This study has also been funded in part by the Brain & Behavior Research Foundation via the 2020 NARSAD Young Investigator Grant #29350 (GV).

