## Supplementary_methods for "Genome-wide association study of treatment resistant depression highlights shared biology with metabolic traits"

**Study Settings**

MGB consists of 2 academic medical centers and 4 community and psychiatric hospitals in Eastern Massachusetts that serve over 6.5 million patients, and electronic health data were extracted from the Mass General Brigham Research Patient Data Registry (RPDR)^19^ and the Enterprise Data Warehouse. VUMC is an academic medical center in Nashville, Tennessee that manages over 2 million patient visits across Tennessee and its neighboring states each year. Its deidentified clinical EHR data is stored in the Synthetic Derivative (SD)^20^. Geisinger Health System is an academic medical center in Danville Pennsylvania and serve over 3 million patients in Pennsylvania. Deidentified electronic health data for consenting patients is extracted and stored by the Geisinger MyCode Community Health Initiative^21^. The Million Veteran Program^22^ study is based on the largest integrated health care system in the United States (Veterans Health Administration; 9 million people) and has over 825,000 US veteran participants with genetic information available for over 650,000 individuals.

**Clinical prediction model of TRD**

Individuals with MDD were identified using International Classification of Diseases, version 9 (ICD-9) codes 311.* (depression not otherwise specified), 296.2* (depressive episode), 296.3* (recurrent depression), and 300.4 (dysthymic disorder) and ICD-10 codes F32.** (depressive episode), F33.** (recurrent depressive disorder), and F34.1 (dysthymic disorder) with * as wildcard digits 0-9.

Structured clinical data included as predictors for the clinical model, are: demographics (age in years, categorical sex [Male, Female, Unknown], categorical race [White, Black, Asian, Hispanic, Other]), area deprivation index (ADI), diagnostic codes (log-transformed counts of historical Clinical Classification Software (CCS) counts)^23^, and medication (log-transformed counts of RxNorm-mapped ingredients). Of note, the VUMC ADI uses six features from the American Community Survey on the census tract level^24^, while MGB ADI includes 21 socioeconomic factors from the census on the zip-code level^25^.

**Genotyping, quality control and GWAS of the MGB sample**

We obtained the recently released genotyping array data for 36,424 subjects in the MGB Biobank. Genotyping of MGB samples was performed using the three versions of the Illumina Multi-Ethnic Global (MEG) array (Illumina, Inc., San Diego, CA): MEGA (N = 4,924; 1,416,020 SNPs), MEGAEX (N = 5,344; 1,741,376 SNPs), and MEG (in 6 batches; N = 26,156; 1,778,953 SNPs) on the human genome build hg19 coordinates. To avoid batch effects, we performed quality control (QC) steps separately for each genotyping batch to remove SNPs with genotype missing rate > 0.05, samples with genotype missing rate > 0.02, and SNPs with differential missing rate > 0.01 between any two batches. Data were then merged across all batches for subsequent QC steps.

As participants in the MGB Biobank are from diverse populations, we inferred their genetic ancestry using principal component analysis (PCA) with the 1000 Genomes Project (1KG) (The 1000 Genomes Project Consortium et al. 2015) as the population reference panel. Specifically, we merged the MGB samples with the 1KG samples and performed PCA based on a list of high-quality, common SNPs (SNP call-rate > 0.98, minor allele frequency (MAF) > 0.05, non-strand-ambiguous, and not in long-range linkage-disequilibrium (LD) regions) after LD pruning. Using the first 6 PCs of the 1KG samples, we then trained a Random Forest classifier that assigned a “super population” label for each biobank sample and retained samples with a prediction probability ≥0.9. This resulted in 26,677 individuals classified as the European (EUR) ancestry, 1,607 Africans (AFR), 1,840 Admixed Americans (AMR), 504 East Asians (EAS), and 297 South Asians (SAS).

Within the EUR ancestry, we removed samples whose reported and genetic sex did not match, those with outlying values of the absolute value of heterozygosity (> 5SD from the mean), and one from each pair of related individuals (IBD > 0.2). SNPs that showed significant batch associations at *P* < 1 × 10^−4^, with a missing rate > 0.02, or an HWE test *P* < 1 × 10^−10^ were also discarded. Next, we imputed genotype dosages for the biobank samples on the Michigan Imputation Server (Minimac4) using the Haplotype Reference Consortium (HRC) reference panel. Lastly, we removed genetic markers with an imputation quality INFO score < 0 .8, MAF < 0.01, a significant deviation from HWE at *P* < 1 × 10^−10^, and a missing rate > 0.02.

TRD posterior probabilities were inverse rank normalized and GWAS analysis was performed using linear regression in PLINK 2.0 with covariates for the top 10 principal components.

**Genotyping, quality control and GWAS of the VUMC sample**

The 98,473 individuals’ genetic data were genotyped by the Illumina Infinium expanded multi-ethnic genotyping array (MEGAEX), which contains 2,038,233 SNPs. SNP quality control steps include excluding SNPs with MAF < 0.005, Hardy-Weinberg equilibrium test P value ≤ 10^-10^ within each self-reported ancestry or call rate <95%. Individuals were removed if they had a mismatch between genetically inferred sex and self-reported sex, excess heterozygosity rate within each self-reported ancestry, missing rate ≥ 0.02, or potentially cross-contaminated samples (proportion IBD > 0.8). 94,369 samples and 887,250 high-quality autosomal SNPs remained.

Ancestry was determined with 1000 Genomes phase 3 (1000GP3) data. 1000 Genomes phase 3 (1000GP3) consists of 2,504 unrelated samples from 5 super populations African (AFR), Admixed American (AMR), East Asian (EAS), European (EUR), South Asian (SAS). 887,250 genotyped autosomal SNPs from the BioVU MEGAEX array was merged with 1000GP3 after removing C/G and A/T SNPs to avoid unresolvable strand mismatches in MEGA samples. Regions with known high LD^30^ (Chr 5 44–51.5 Mb, Chr 6 25–33.5 Mb, Chr 8 8-12Mb, Chr 11 45–57 Mb) were excluded and the common variants were then pruned (r2 < 0.05) using PLINK 1.9^31^ (–indep-pairwise 1000 50 0.05) to yield 71,339 SNPs in relative linkage equilibrium for ancestry analyses. Principal components (PCs) were generated using flashpca version 2.0. By using K nearest neighbors (KNN, k = 50) clustering, we inferred MEGA samples' ancestries. We treated 1000GP3 samples' PCs as the training set and MEGA samples' PCs as the test set. For each individual within the MEGA sample, we calculated its Euclidean distance to all reference individuals based on the 2 leading PCs and then identified the 50 nearest individuals. If at least 90% of the closest individuals were from the same super population, we inferred that the MEGA individuals belonged to that super population. Individuals not surpassing that threshold were considered admixed. Among the 94,369 MEGA individuals, 91,141 (96.6%) were assigned to a homogeneous super-population, with the following breakdown: AFR=14,176, AMR=1,152, EUR=74,612, EAS=762, SAS=439. The total set of individuals considered admixed was 3,228. A subset of individuals of EUR ancestries (MEGA-EUR) were selected for further analysis. All 94,369 samples were imputed on the Michigan Imputation Server v.1.2.4 using Eagle (V2.4.1) for phasing, Minimac4 for imputation and the Haplotype Reference Consortium (HRC) reference v1.1 panel in build GRCh37 as reference. Genotype probabilities were converted to hard-call genotypes using PLINK2 (hard-call ≥ 0.1). SNPs were filtered with imputation info score in any of the batches < 0.8 missing genotype rate > 0.02, or multi-allelic states (>2). Within EUR super populations, SNPs with MAF < 0.005 and Hardy-Weinberg equilibrium test P value < 1 × 10^-10^ were excluded.

TRD posterior probabilities were inverse rank normalized to a mean of 0 and a standard deviation of 1. Quantitative GWAS of TRD posterior probabilities was performed using covariates of sex, age and PC1-PC20 (22 covariates) using Regenie v1.0.7, a computationally efficient method of whole genome regression modeling for genome-wide association analyses^32^ (**Figure 1C**). Default settings of block size 200 and 20 threads were used.

**Genotyping, quality control and GWAS of the GHS sample**

The GHS sample was genotyped under the DiscovEHR Collaboration between Geisinger and the Regeneron Genetics Center. Patient-participants were genotyped with (a) the Illumina Human OmniExpress Exome array or (b) the Illumina Global Screening Array. Therefore, downstream quality control and imputation was performed separately per array, and all association analyses controlled for an array dummy. The genotype data was imputed to the Haplotype Reference Consortium by Regeneron with the Minimac4 protocol [HRC Ref: McCarthy et al., 2016, Nat Genetic]. Pre-imputation filtering included removing SNPs with (1) MAF less than 1%, (3) HWE-equilibrium *P* less than 1×10^–15^, and missingness greater than 1%. Samples were included if they satisfied (1) autosomal missingness less than 2%, (2) no excessive PC-adjusted heterozygosity, (3) no X-chromosome mismatch with reported sex, (4) no sex mismatch based on an F_X_ less than 0.5 for self-reported males, and F_X_ greater than 0.5 for self-reported females.

Post-imputation quality control consisted of identifying (1) a genetically homogenous cluster of European-ancestry individuals (to safeguard against population stratification in addition to including genetic PCs in GWAS), and (2) pre-GWAS SNP filtering of the imputed genotypes. With respect to (1), the 1000 Genomes phase 3 version 5 reference panel was used to generate genetic PCs to infer ancestry in the Geisinger population. We identified a set of 36,647 high-quality and near-independent SNPs for principal component analysis (PCA) using the 1000 Genomes data. The Geisinger samples were then projected on the first four PCs, which clearly separated a tight European-ancestry cluster from the other major continental ancestral groups, in addition to utilizing self-reports on “White” and “Not Hispanic or Latino” background. At this stage, 124,467 ancestrally homogenous samples were subjected to iterative principal component analysis using 69,862 quality-controlled and LD-pruned SNPs, resulting in a final sample of 122,370 patient-participants that were eligible for inclusion in this study. A final round of PCA was conducted to generate PCs for inclusion as control variables in GWAS.

Concerning (2), pre-GWAS SNP filtering consisted of removing SNPs (a) with MAF less than 0.1%, (b) with imputation INFO less than 0.2, (c) with average maximum posterior call less than 0.9, and (d) keeping only biallelic SNPs (dropping multi-allelic and insertions/deletions). Then, GWAS was conducted on the quantitative TRD phenotype as leave-one-chromosome-out (LOCO) linear mixed models by using the SAIGE software (v.0.44.3). Covariates were age, sex, array dummy, blood vs. saliva dummy, and 20 genetic PCs. The genetic variance component consisted of a lightly LD-pruned set of 447,896 SNPs that passed all aforementioned quality-control criteria, as well as dosage certainty greater than 0.8 and hard-call missingness less than 0.05. The association analysis itself was conducted on imputed dosages and not hard calls (while the variance component requires hard calls). The final GWAS sample size in GHS was 39,353.

**Genotyping, quality control and GWAS of the MVP sample**

MVP Genotyping, initial quality control, and imputation is handled by the MVP Data Team as previously described^29^. This study used MVP release 4, which includes genotyping data from just over 650,000 individuals. DNA was extracted from blood of MVP participants during enrollment, and genotyping was performed with MVP 1.0 custom Genotyping array^29^ and genotype calling with APT software 2.11.3. Genotype quality control metrics include among others: sex check, rare heterozygous adjustment, plate normalization and sample labeling check. Ancestry-specific principal component analysis was performed with PLINK 2.0^30^ and relatedness (kinship coefficient) was calculated using KING 2.1^31^. Ancestry was determined by HARE^32^ (genetically inferred ancestry was based on top 30 principal components and ADMIXTURE^33^) and genotypes were phased with SHAPEIT4^34^ version 4.1.3. Pre-imputation QC removed variants that had high missingness (> 20%), were monomorphic, or had high differences in frequency between batches. Genotypes were imputed with 1000 Genomes^35^ (except for African individuals, which were imputed with Sanger Institute African Genome Resources Panel^36^ using Minimac4^37^. Then, we performed sample-level quality control which: a) retained samples with sample call rates > 98.5%, b) retained samples with sample heterozygosity rates which deviate 4SD or less from the samples’ heterozygosity rate mean and c) removed related samples and samples with cryptic relationship with a kinship coefficient cut-off of ≥ 0.0884 (first samples with multiple relationships were removed; then samples with the highest missingness rates were removed from the remaining relationship pairs). Variant-level filtering was performed by retaining variants with MAF > 0.005, effective minor allele count ≥ 30, variant call rates > 95%, Hardy-Weinberg equilibrium test p value > 5 × 10^-8^, and imputation R^2^ > 0.4.

TRD posterior probabilities were inverse rank normalized to a mean of 0 and a standard deviation of 1. GWAS analysis was conducted employing a linear regression in PLINK 2.0 using covariates for age, sex, and top 20 principal components.

**Conditional GWAS using mtCOJO**

mtCOJO analysis was performed on the TRD meta-analyses as the outcome traits with the GIANT European ancestries GWAS summary statistics^36^ as the exposure trait since mtCOJO requires an ancestry-matched LD reference panel. In the selection of SNPs as instruments, independence was defined as SNPs more than 1 megabase (Mb) apart or with an LD r2 value < 0.05 based on the 1000 Genomes Project Phase 3 European reference panel^37^.

**Supplementary Results**

**Validation of ECT cases as a TRD surrogate**

Among the VUMC MDD cohort, 615 patients had direct documentation of TRD in their clinical record including 105 or 47% of the ECT cases representing a 182-fold increase of direct TRD documentation compared to the rest of the MDD cohort. The MGB dataset omits narrative clinical notes from McLean Hospital prior to 2017, which would tend to underestimate documentation of TRD given the greater proportion of inpatient care delivered there. Nevertheless, 266 MDD patients with documentation of TRD were identified including 41 or 15% of those with ECT which was a 58-fold increase of direct TRD documentation compared to the remaining MDD cohort.

In addition to gathering additional evidence of TRD among ECT cases, we also tested for phenotypes known to associate with TRD, such as increased suicidality and higher burden of comorbid psychiatric illnesses. We performed a phenome-wide association study (PheWAS) to test the relationship of ECT with other phenotypes (see Methods). PheWAS association results in the VUMC and MGB cohorts were highly correlated (r = 0.70. P = 4.19x10^-54^), with suicidality being the most significantly associated phenotype in both VUMC and MGB (VUMC: beta = 3.62, SE = 0.15, P = 2.67x10^-128^; MGB: beta = 2.57, SE = 0.18, P = 2.58x10^-46^). Other significantly associated phenotypes include psychiatric diseases like major depressive disorder, generalized anxiety disorder and other suicide-related traits (**Supplementary Figures 1-2**).
