## Supplementary_figures for "Genome-wide association study of treatment resistant depression highlights shared biology with metabolic traits"

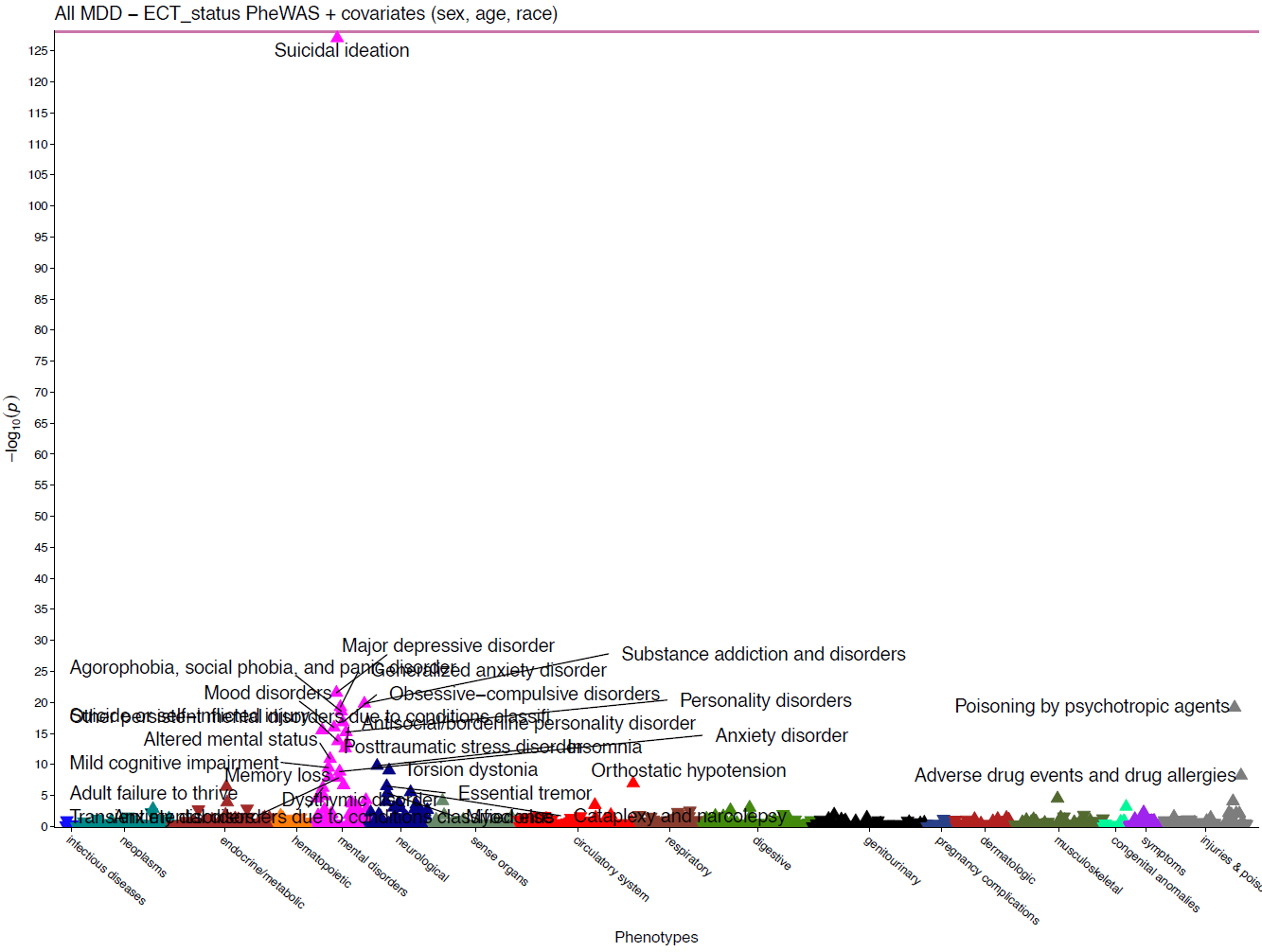

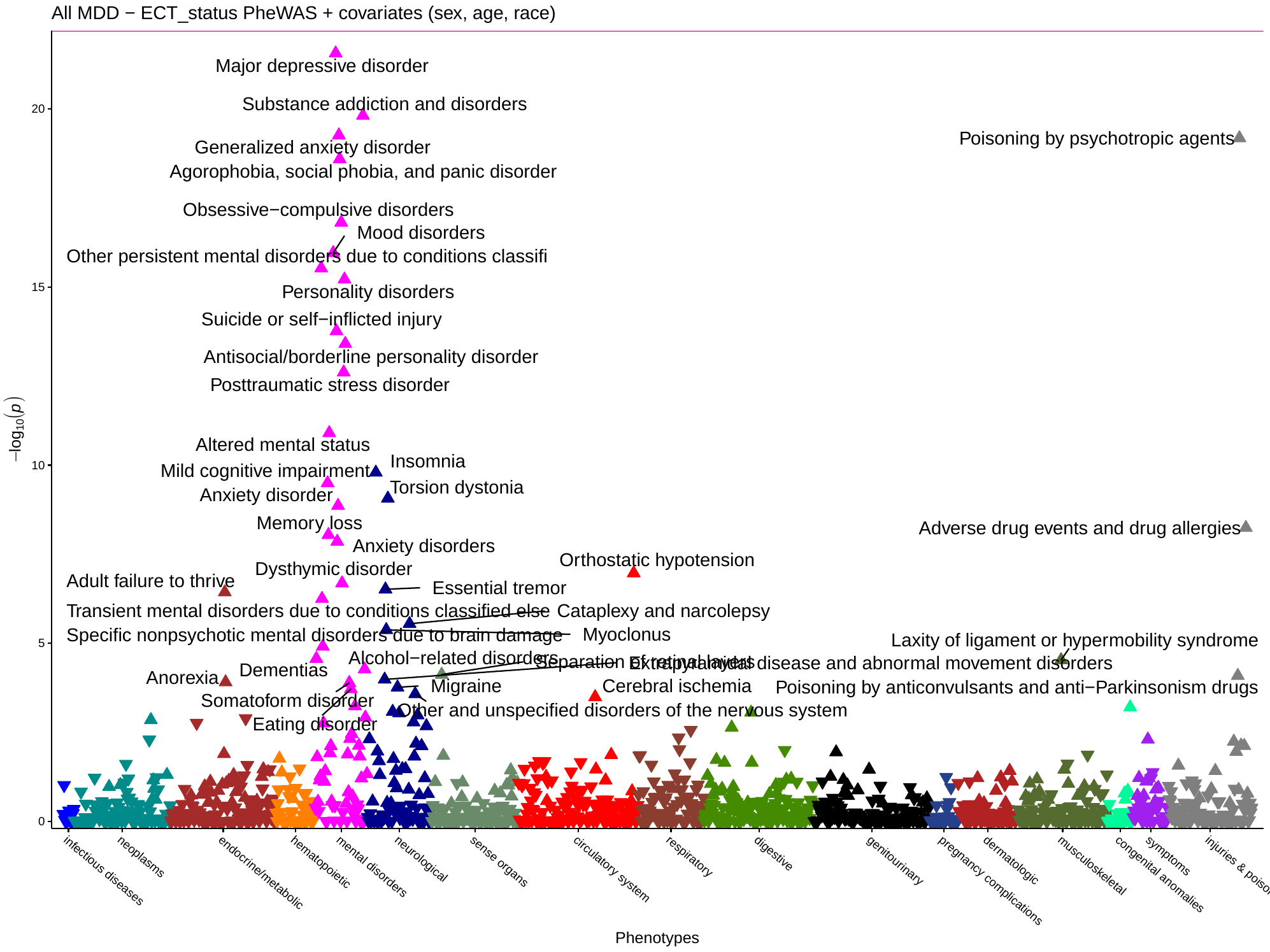

**Supplementary Figure 1: Phenome-wide association study of ECT CPT code among all MDD patients in VUMC.**

Phecodes with counts over 100 were included for analysis. Covariates of the regression included sex, age, and race. In the second plot, the strongest association, suicidal ideation (p=2.67x10^-128^) was omitted for scale.

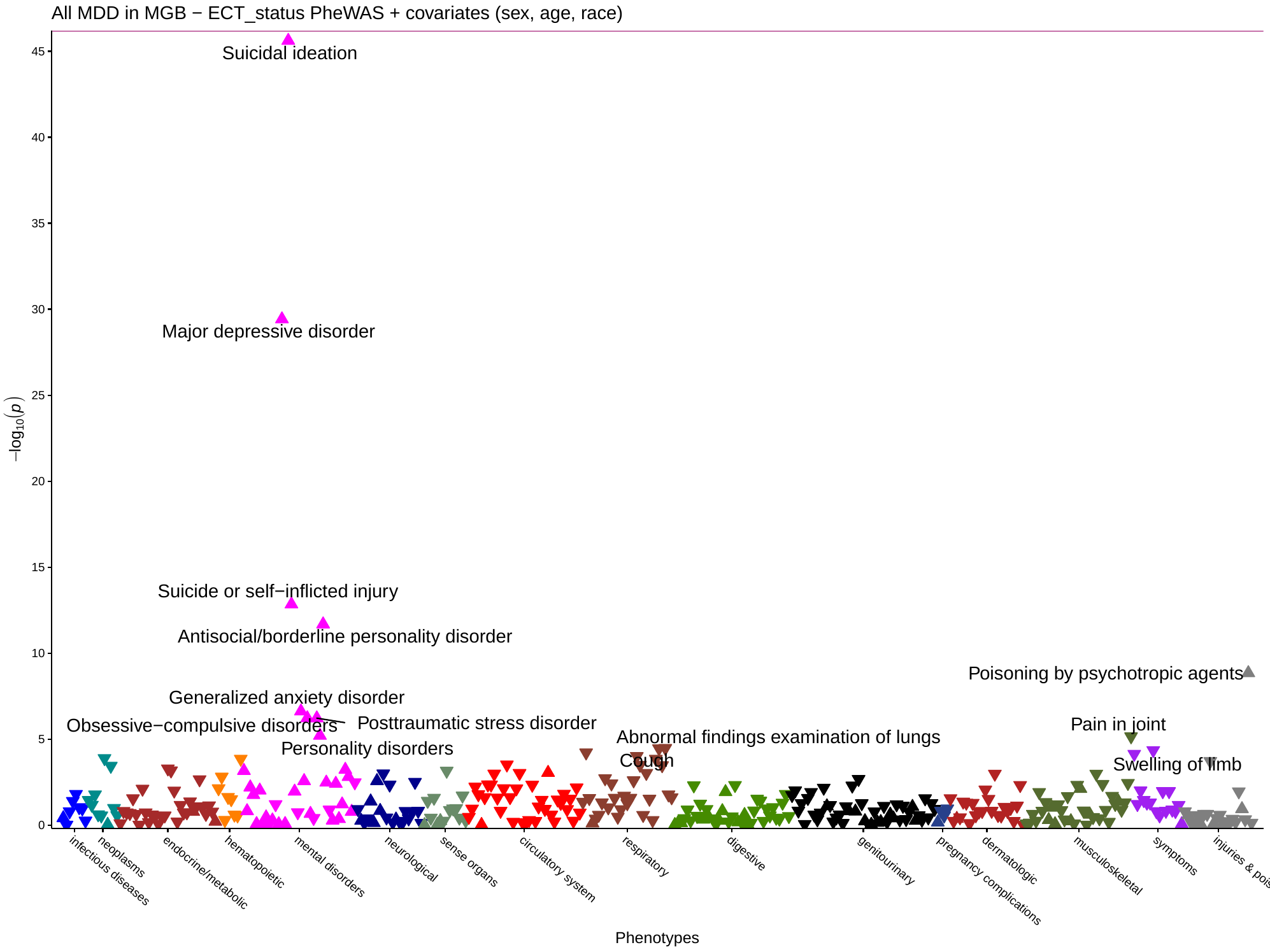

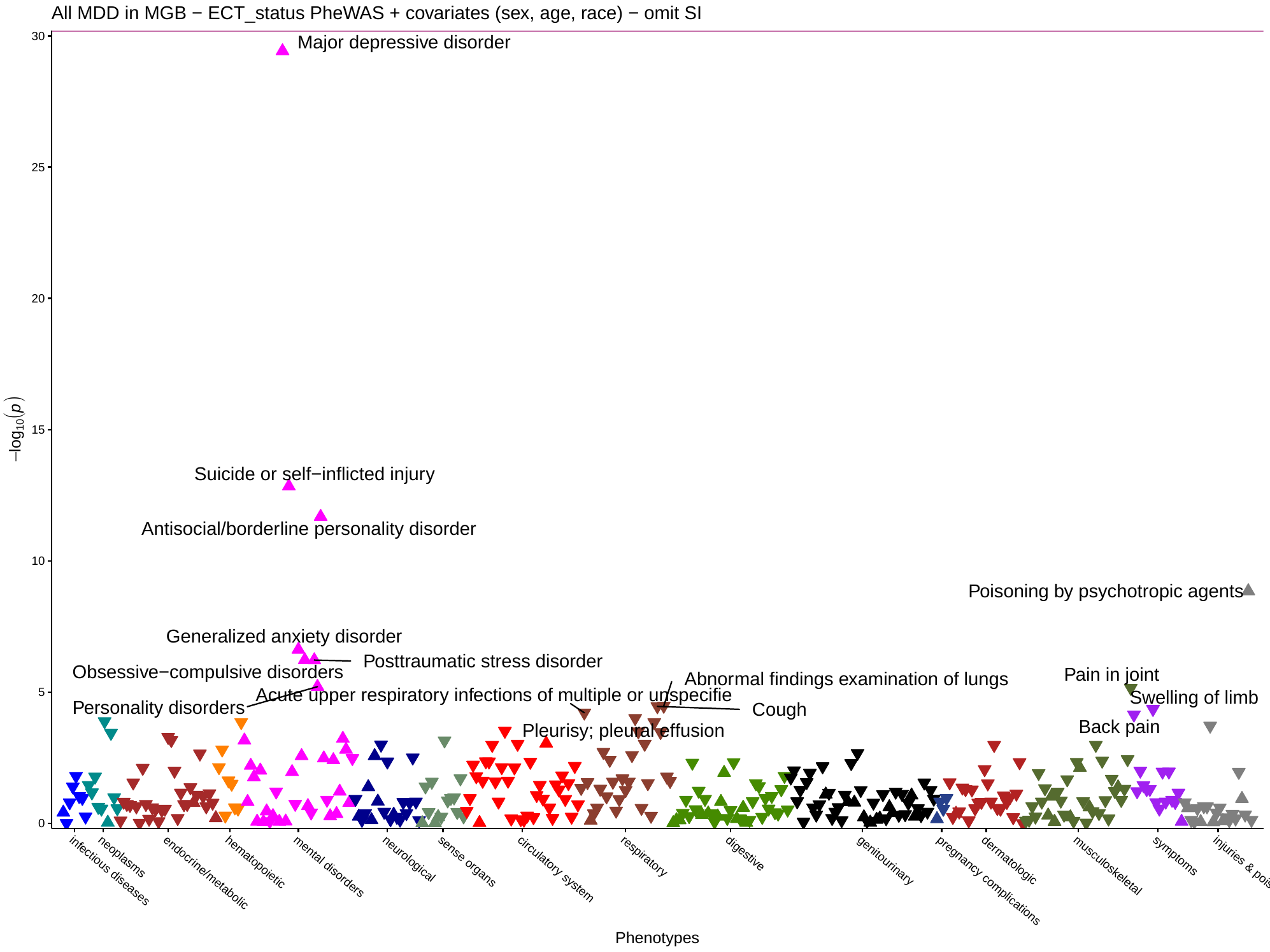

**Supplementary Figure 2: Phenome-wide association study of ECT CPT code among all MDD patients in MGB.**

Phecodes with counts over 100 were included for analysis. Covariates of the regression included sex, age, and race. In the second plot, the strongest association, suicidal ideation (p=2.39x10^-46^) was omitted for scale.

**Supplementary Figure 3:** **Genetic correlations of MGB TRD meta-analysis GWAS with psychiatric and non-psychiatric traits before and after conditioning for BMI.**

Unfilled points indicate genetic correlations that did not pass the Bonferroni-corrected significance threshold P<1.72x10^-3^ (29 traits tested). Error bars represent the standard error. P values indicate significant differences in genetic correlation after conditioning, that pass the Bonferroni correction. Bolded traits show significant differences in genetic correlations between the TRD meta-analysis before and after BMI conditioning with mtCOJO. BMI-body mass index, ADHD-attention-deficit/hyperactivity disorder, OCD-obsessive compulsive disorder, PTSD-post-traumatic stress disorder, AUDIT-C-Alcohol Use Disorders Identification Test-C (measure of quantity of alcohol consumption), AUDIT-P- measure of problematic consequences of drinking, AUDIT-T-total score of AUDIT.

**Supplementary Figure 4:** **Genetic correlations of VUMC TRD meta-analysis GWAS with psychiatric and non-psychiatric traits before and after conditioning for BMI.**

Unfilled points indicate genetic correlations that did not pass the Bonferroni-corrected significance threshold P<1.72x10^-3^ (29 traits tested). Error bars represent the standard error. P values indicate significant differences in genetic correlation after conditioning, that pass the Bonferroni correction. Bolded traits show significant differences in genetic correlations between the TRD meta-analysis before and after BMI conditioning with mtCOJO. BMI-body mass index, ADHD-attention-deficit/hyperactivity disorder, OCD-obsessive compulsive disorder, PTSD-post-traumatic stress disorder, AUDIT-C-Alcohol Use Disorders Identification Test-C (measure of quantity of alcohol consumption), AUDIT-P- measure of problematic consequences of drinking, AUDIT-T-total score of AUDIT.
